# FounderRare: A Novel Statistical Package to Identify Rare Variants in Complex Diseases

**DOI:** 10.1101/2025.09.11.25335516

**Authors:** Samir Oubninte, Simon Girard, Claudia Moreau, Alexandre Bureau

## Abstract

`FounderRare` is a statistical package designed for the identification of rare variants in complex diseases. It leverages population genealogies, including those shaped by founder effects and identity-by-descent (IBD) segments, distinguishing it from other tools. This paper serves as a theoretical guide to the `FounderRare` package, illustrating its operations and capabilities. The package implements an IBD-based approach that computes the number of copies, among affected individuals, of the shared haplotype within genomic regions. The genome is partitioned into regions, within which clusters of affected individuals sharing specific IBD segments are identified. Statistical tests, denoted as *S*_*msg*_ and *S*_*all*_, are included to evaluate the enrichment of IBD sharing among affected individuals. These tests rely on simulations of the null distribution and are designed to identify causal regions in the absence of control samples. `FounderRare` is optimized for cohorts comprising several thousand individuals—sample sizes typically required to achieve sufficient statistical power in rare variant analyses. By utilizing genotype array data, this tool enables cost-effective analysis at scale for researchers investigating complex diseases. It aids in pinpointing genomic regions likely to harbor rare variants, thereby contributing to a deeper understanding of the underlying genetic structure.

FounderRare, R package, rare variants, complex diseases, population genealogy, identical-by-descent (IBD) segments

## 1 Introduction

Complex diseases are shaped by an interplay of genetic and environmental elements. Despite the progress in genome-wide association studies (GWAS), a significant portion of the genetic variance of these diseases remains elusive, leading to the so-called “missing heritability” problem [1, 2]. One potential explanation for this missing heritability could be the role of rare variants [3], which are hypothesized to have larger effects but lower frequencies compared to common variants. However, the detection of these rare variants poses a significant challenge due to their low statistical power, high heterogeneity, and limited availability of large sample sizes of whole genome sequences.

To address these challenges, we introduce a statistical method that leverages both genealogical and genomic data from a population, including those with a founder effect, to pinpoint rare variants associated with complex diseases [4]. A population with a founder effect is a group that descends from a limited number of ancestors who possess a subset of the genetic diversity of the original population. Owing to the decreased genetic diversity and increased relatedness, populations with a founder effect are a rich source of rare variants. These rare variants are often shared among relatives and are located on Identical by Descent (IBD) segments, which are genomic regions inherited from a common ancestor without recombination. By tracking these IBD segments within a population, we can deduce recent variants that are passed down the genealogical tree and examine their association with disease phenotypes [7]. This approach provides a promising avenue to uncover part of the hidden heritability of complex diseases and advance our understanding of their genetic architecture.

We have developed `FounderRare`, an R package that encapsulates our statistical methodology and a statistical measure of the most shared haplotype in a synthetic genomic region among affected individuals, known as *S*_*msg*_. This measure is used in the approach we developed to identify rare variants in populations with a founder effect [4]. `FounderRare` accepts IBD clusters as input and subsequently partitions the genome into synthetic genomic regions (SGs), guided by the IBD structure. It then identifies distinct clusters of affected individuals who share a specific SG. Following this, the value of statistical measures like *S*_*msg*_ is computed.

The effectiveness of `FounderRare` was evaluated using simulated and real data sets in a separate study [4]. The simulation of whole genome transmission in a Quebec founder genealogy (sourced from the BALSAC population database, website: https://balsac.uqac.ca/) was performed using msprime [9]. Our approach was then compared with alternative strategies such as the Generalized Linear Mixed Model Association Test (GMMAT) [5] and the *S*_*all*_ measure (an adapted version of *S*_*all*_ proposed by Whittemore [6]). `FounderRare` can compute this adapted measure of *S*_*all*_. The results indicated that *S*_*msg*_ implemented in `FounderRare` can be more powerful and robust than certain competing IBD segment test statistics, especially in identifying regions that contain a single causal variant.

The `FounderRare` package is complemented by a comprehensive guide that explains its functionalities and presents typical examples of its output. The guide delves into the package installation process, detailing the necessary prerequisites and dependencies. It also provides a catalog of the functions embedded in the package, offering insights into their usage, thereby allowing for package adaptability for future research. Furthermore, the guide demonstrates the application of `FounderRare` on real datasets. One such dataset, named IBDClusters, is included as an example. This dataset is an output generated by Dash [7], a tool designed to infer clusters of individuals who are likely to share a single haplotype.

## 2 Methods and Implementation

The `FounderRare` package is a tool designed to detect rare variants associated with complex diseases within a population with a founder effect, using IBD segments as a proxy. This package offers a set of functions that streamline the analysis process, including the creation of synthetic genomic regions, the examination of IBD structures, and the calculation of the statistic *S*_*msg*_ for each SG.

To effectively utilize the `FounderRare` package, it is recommended to acquire IBD clusters from a population with a founder effect. These necessary clusters are used in the output format of the Dash software. We adopted an affected-only strategy, which allowed us to focus on those individuals in the population who exhibited disease traits. Upon identifying IBD clusters using Dash or any other suitable tool, we follow the recommended workflow:

1. **Defining Initial Genomic Ranges for SG and Sliding-SG:** The SG is defined by its ***Start*** and ***width***. The start is the smallest cluster start position on the chromosome. The width is minL *×* 1000, where minL (default: 400) is the initial length in kilobases, giving an initial width of 400 Kbp. The Sliding-SG is constructed similarly, using a fraction of this width and a different position (see Figure 1 and the tutorial). For each SG, an associated Sliding-SG is created, overlapping the SG by half its length. Optional parameters allow users to adjust both SG and Sliding-SG for specific analyses.
2. **Analysis of IBD Structure and Generation of the Subsequent SG:** This procedure involves identifying and comparing IBD structures between two SGs, specifically an SG and its corresponding Sliding-SG. The process begins with the function `IBD structure()`, which determines the IBD structure, defined as the set of clusters satisfying a specified overlap criterion within a given genomic range. This is controlled by an optional overlap cutoff specifying the minimum proportion of overlap between clusters and the SG range required for inclusion (default: 0.5). The resulting structures are then compared. If the SG and Sliding-SG share the same IBD structure, the SG is extended to the end of the Sliding-SG, and the output includes the extended SG genomic range (returned as a `GRanges` object, following Bioconductor conventions) and its IBD structure (see Table 1). If the structures differ, indicating distinct patterns, a new SG and its associated Sliding-SG are created. The new SG starts at the end of the current SG, with an initial length of 400 Kbp (default value of ***minL***).
3. **Ensuring Disjointedness of IBD Structures:** The `FounderRare` package applies the Tunnel Boring Machine (TBM) principle—an iterative approach that progressively generates or enlarges SGs based on their IBD structures until the end of the chromosome is reached. This ensures that the SG under examination encompasses the farthest of all cluster ends. Once this step is complete, the extraction of disjoint clusters begins. This involves reviewing each SG to determine whether its IBD structure contains overlapping (i.e., non-disjoint) clusters (see Table 2). If overlaps are detected, two strategies are available to achieve disjointness. The recommended approach is to retain the clusters with the largest number of individuals among the overlapping clusters [4]. Alternatively, overlapping clusters can be merged.
4. **Computation of Shared SG Statistic:** The value of IBS sharing statistics, such as *S*_*msg*_, involves computing the value of 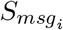 for every SG using Equation 1. To account for varying sample sizes, the *S*_*msg*_ statistic can be normalized by the number of affected individuals.

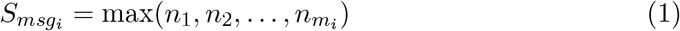

where *m*_*i*_ represents the number of disjoint clusters in *SG*_*i*_, and 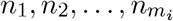 represent the number of haplotypes in each cluster (*i* = 1, …, *I* = number of constructed SGs).

**Figure 1.**
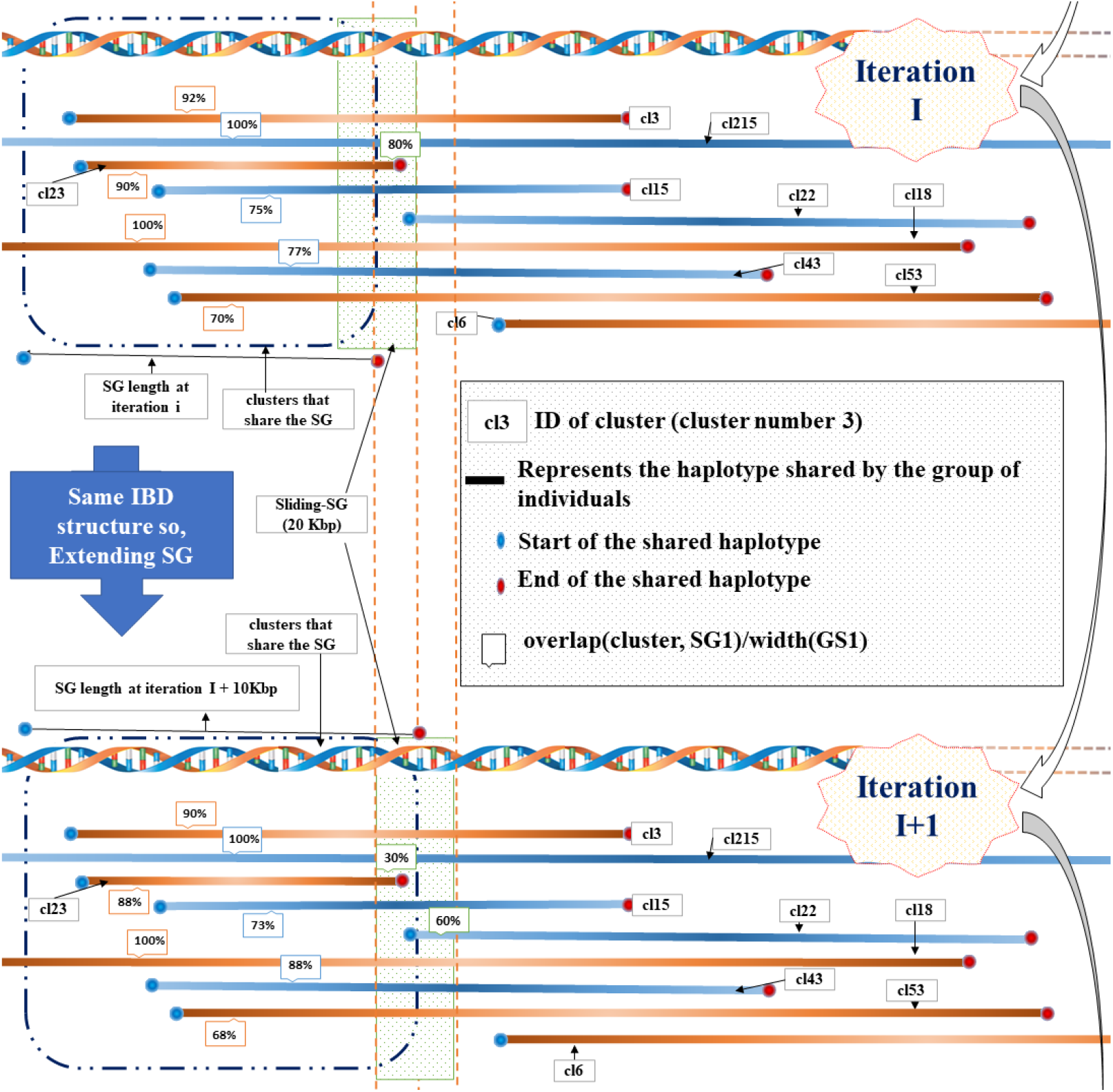
Overview of SG Construction: This schematic illustrates the process of identifying and constructing SGs across two successive iterations (Iteration I and Iteration I+1). Horizontal lines, displayed in blue and brown, represent haplotypes shared among clusters of individuals, labeled with unique identifiers (e.g., cl3, cl15, cl22). The lengths of these lines vary, reflecting the diversity of shared haplotypes. Dashed black lines connect clusters that share identical SG start and end points. Percentages along the haplotype segments indicate the proportion of overlap with either the cluster SG or the sliding-SG (represented by the green box labeled “Sliding-SG”). At the bottom of the diagram, SG length is measured, demonstrating that segment lengths can vary across iterations. A vertical brown dashed line highlights an example where the SG length increases by 10 Kbp from Iteration I to Iteration I+1, attributed to the IBD structure of the SG being similar to that of the sliding-SG. Specifically, clusters with SG overlap ≥ 50% also exhibit sliding-SG overlap ≥ 50%. In subsequent iterations, the algorithm is expected to generate a new SG. *This figure is reproduced from Oubninte et al*.*[4]*

**Table 1:**
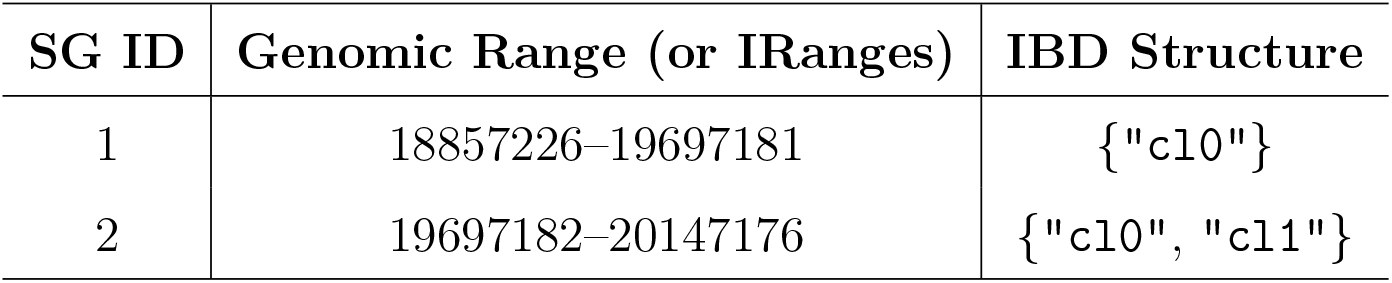
SG Data Structure: Data structure for the first two SGs. Each SG is represented as a list containing two components: (i) the genomic range (as a GRanges or IRanges object) and (ii) the clusters included in its IBD structure. One cluster—cl0—appears in overlap between SG 1 and 2.

**Table 2:** Disjoint Clusters within SGs: Example of the output generated by the `TBMD()` function for the first two SGs. Each SG is represented by its genomic range and the corresponding sets of disjoint clusters—namely the input clusters cl0 and cl1, reformatted according to the SG structure. Each cluster consists of individuals sharing a haplotype; the suffix (e.g., .2) indicates the haplotype index for that individual.

| SG ID | Genomic Range | Disjoint Clusters |
| --- | --- | --- |
| 1 | 18857226–19697181 | {"2349.2", "2739.2"} |
| 2 | 19697182–20147176 | {"2125.1", "2737.1", "2349.1"}<br>{"2349.2", "2739.2"} |

Additionally, the `FounderRare` package offers the functionality to preserve SGs with their original IBD structure, as well as the IBD structure with disjoint clusters. This dataset may serve as a foundation for developing new statistical measures that extend or improve upon the concept of the *S*_*msg*_ statistic. Essentially, the `FounderRare` package is a toolkit that has been tested for the investigation of rare variants in complex diseases within a population with a founder effect. We have made available a separate guide on *GitHub* that details the necessary steps to download, install, and set up the `FounderRare` package. This guide includes comprehensive code examples to aid first-time users.

## 3 Practical Application: Harnessing the Power of the FounderRare Package

In this section, we illustrate the application of the `FounderRare` package to identify rare variants in complex diseases by analyzing IBD structures. We provide a step-by-step guide on how to use the functions detailed in the package’s documentation for basic usage. For more comprehensive information, including additional functions and parameters that can be customized to the user’s needs, please refer to the accompanying vignette of the package. It provides detailed information about the function’s purpose and usage, thereby enhancing the user’s grasp of its application.

1. **Downloading and Installing** `FounderRare` **Package:** To successfully download and install the `FounderRare` package, adhere to the guidelines provided in the vignette document. The required libraries, including `GenomicRanges`, `FounderRare`, `dplyr`, and `tidyr`, must be loaded as they furnish the necessary tools for effective data manipulation and analysis.
2. **Inferring Synthetic Genomic Regions:** The inference of SGs can be achieved using the TBM() function, which constructs SGs based on the TBM principle. The function dynamically creates or expands SGs based on their Identity by Descent structures. It accepts several parameters, including IBD clusters (IBDClusters), and various overlap and length parameters. This process involves the utilization of numerous other functions provided by the package. Below is an example of R code to infer SGs using the recommended (default) values:

~~~
# An example of Identity by Descent clusters information
           is loaded from the package.
# For details on how to read IBD clusters from Dash output files,
# please refer to the package’s vignette.
data(“IBDClusters”)
# Columns containing only NA values are removed
     from the IBDClusters data frame.
IBDClusters = IBDClusters[,
           colSums(is.na(IBDClusters)) != nrow(IBDClusters)]
# The TBM function is called to infer SGs.
     It uses the IBDClusters
       and chromosome information,
# with default values for other parameters.
 SGList = TBM(IBDClusters, chr = 22)
SGList # This will display the information
          of the created SG on the console.
# The inferred SGs, along with their IBD structure,
             can be saved in an RData file.
# The filename should be specified by the user.
# We recommend that you customize the file name and specify the path.
saveRDS(SGList, file = paste0(path, “SGList_chr”, chr, “.RData”))
~~~

3 **Disjointness in IBD Structures of SGs and** *S*_*msg*_ **Computing:** The `TBMD()` function is designed to transform the non-disjoint clusters present in the output list of SGs (SGList) from the `TBM()` function into a refined output comprising disjoint clusters. It sequentially examines each SG to determine whether the IBD structure within the SG encompasses non-disjoint clusters. By default, it employs the `Disj Clst SG Bigger()` function, which converts the IBD structure into disjoint clusters by retaining the clusters with the most individuals among overlapping clusters.

~~~
# The TBMD() function is used to infer the SG with disjoint clusters.
SGListWithDisjCls <-TBMD(SGList, IBDClusters)
SGListWithDisjCls # This will display the details of
                    the generated SG in the console.
# The output can be saved into an RData file.
saveRDS(SGListWithDisjCls,
            file = paste0(path, “SGListWithDisjCls_chr”, chr, “.RData”))
# The Smsg statistic is computed using the ‘SGListWithDisjCls’ output. outSmsg <-Smsg(SGListWithDisjCls)
# The output can be viewed by using the print() function
print(outSmsg)
#> chr SG cd Start End Smsg
#> 1 22 SG1 cd1 18857226 19697181 2
#> 2 22 SG2 cd1 19697182 20147176 3
# The computed Smsg statistic can be written to a csv file.
write.table(outSmsg, paste0(SGListWithDisjCls, “.csv”),
          sep = “,”, quote = FALSE, row.names = FALSE,
                         col.names = TRUE)
~~~

In the final step, the distribution of *S*_msg_ under the null hypothesis (H0) is needed. In the absence of controls, Oubninte et al.[4] used the affected subjects’ genealogy to simulate neutral genetic sequence data under a coalescent model using Msprime. This simulated whole-genome sequencing data is then used to infer clusters. Following this, steps 2 and 3 are applied to each replicate to generate the null distribution for the statistic. *S*_msg_ is subsequently calculated for the real data. The causal regions that potentially harbor a rare variant are identified as those where the value of *S*_msg_ exceeds a predetermined quantile of the distribution of *S*_msg_ under H0.

## 4 Discussion

The `FounderRare` package equips researchers with advanced tools to investigate the genetic architecture of complex diseases. This package is specifically designed to identify rare genetic variants that contribute to the etiology of these diseases. Noteworthy features include its scalability, possibility to use genotype array data, focus on affected individuals, ability to infer synthetic genomic regions and capacity for rapid chromosome scanning using moderate resources: generating the SGs is a process taking several minutes per chromosome, while ensuring disjointness of IBD structure and computing *S*_msg_ takes merely a fraction of a second. An additional advantage of this package lies in its ability to recode the outputs of the synthetic genomic regions into a data frame. This feature enables the application of a diverse array of statistical methods specifically designed for common variants, positioning the `FounderRare` package as a beneficial resource for researchers in the field.

## Data Availability

All data produced in the present study are available upon reasonable request to the authors

## Acknowledgments

Example data used for illustration are taken from the Eastern Quebec Schizophrenia and Bipolar Disorder Study. Signed consent was obtained from all participants or from the parents for participants under 18 years of age for collection of all data analyzed here, under the supervision of the University-affiliated neuroscience and mental health ethics committee. We thank M. Maziade (Laval University) for access to data from this study. We are grateful to all participants who enabled this study by contributing their DNA and to the participants who provided family information enabling the reconstruction of their genealogy.

## Code Availability

Code to run `FounderRare` is available on GitHub: https://github.com/oubninte/run_FounderRare. Information about the `FounderRare` package and its installation is available on https://github.com/oubninte/FounderRare.

## Funding

- This work was funded by Natural Sciences and Engineering Research Council of Canada (NSERC) and Canadian Institutes of Health Research (CIHR)
- Most of the data analyses were performed on computing resources from Compute Canada (now the Digital Research Alliance of Canada).

## Notes

### Competing Interest Statement

The authors have declared no competing interest.

### Author Declarations

Comite d'ethique de la recherche en Neurosciences et sante mentale of the Centre integre universitaire en sante et services sociaux de la Capitale-Nationale gave ethical approval for this work.

### Summary of Updates

This version of the manuscript has been revised to correct an issue in the reference list. One of the cited references was missing and had been incorrectly displayed. The reference has now been properly updated. No other changes have been made to the manuscript.

